# Improving Genetic Association Studies with a Novel Methodology that Unveils the Hidden Complexity of All-Cause Heart Failure

**DOI:** 10.1101/2023.08.02.23293567

**Authors:** John T. Gregg, Blanca E. Himes, Folkert W. Asselbergs, Jason H. Moore

## Abstract

**Motivation:** Genome-Wide Association Studies (GWAS) commonly assume phenotypic and genetic homogeneity that is not present in complex conditions. We designed Transformative Regression Analysis of Combined Effects (TRACE), a GWAS methodology that better accounts for clinical phenotype heterogeneity and identifies gene-by-environment (GxE) interactions. We demonstrated with UK Biobank (UKB) data that TRACE increased the variance explained in All-Cause Heart Failure (AHF) via the discovery of novel single nucleotide polymorphism (SNP) and SNP-by-environment (i.e. GxE) interaction associations. First, we transformed 312 AHF-related ICD10 codes (including AHF) into continuous low-dimensional features (i.e., latent phenotypes) for a more nuanced disease representation. Then, we ran a standard GWAS on our latent phenotypes to discover main effects and identified GxE interactions with target encoding. Genes near associated SNPs subsequently underwent enrichment analysis to explore potential functional mechanisms underlying associations. Latent phenotypes were regressed against their SNP hits and the estimated latent phenotype values were used to measure the amount of AHF variance explained.

**Results:** Our method identified over 100 main GWAS effects that were consistent with prior studies and hundreds of novel gene-by-smoking interactions, which collectively accounted for approximately 10% of AHF variance. This represents an improvement over traditional GWAS whose results account for a negligible proportion of AHF variance. Enrichment analyses suggested that hundreds of miRNAs mediated the SNP effect on various AHF-related biological pathways. The TRACE framework can be applied to decode the genetics of other complex diseases.

**Availability:** All code is available at https://github.com/EpistasisLab/latent_phenotype_project

## Introduction

AHF is a complex trait that results from various underlying conditions, each influenced by multiple genetic and environmental factors, some of which are overlapping. The genetic contribution to these conditions varies: heritability estimates for both left ventricular and congestive heart failure are approximately 0.3 [1, 2], while coronary heart disease (CHD) heritability is 0.5 [3]. Additionally, plaque displays a heritability of 0.8 when calcified and nearly 0 otherwise [4], which adds heterogeneity to CHD diagnoses without categorized arterial plaque. Thus, the distinct but outwardly similar biological states that are classified as AHF arise from genetic, GxE, and epistatic effects that may apply to each state differently. Accounting for this complexity is key to resolving the missing heritability problem, where GWAS-measured heart failure heritability falls short of twin-study estimates [5]. This problem with unexamined heterogeneity worsens significantly with larger GWAS samples [6].

The problems above are examples of unmeasured biological variation, which is a likely reason that AHF has so few GWAS SNP hits [7]. More generally, unmeasured biological variation of a phenotype refers to biochemical or functional differences between instances of the same phenotype. This variation may intentionally or unintentionally be unmeasured, but we assume that failure to measure this biological variation weakens GWAS associations with the phenotype of interest. Conversely, a homogeneous phenotype is one caused by a unique biological state with no such unmeasured variation. GWAS would ideally be conducted against homogeneous phenotypes, but this is often infeasible for complex phenotypes like AHF because indistinguishable outward symptoms often have different genetic causes.

When a presumed homogenous phenotype contains such unmeasured biological variation, we refer to this as phenotypic heterogeneity. ICD codes contain substantial phenotypic heterogeneity because they primarily document billable diagnoses rather than precise biological conditions, the consequences of which are illustrated by a study on heart failure ICD codes’ internal consistency. Two cardiologists conducted a detailed abstraction and validation of 705 generalized heart failure (any ICD-9CM 428 code) hospitalizations with curated medical records, and they agreed on only 75 percent of classified cases [8]. Furthermore, most heart failure cases remain undiagnosed until hospitalization [9], suggesting that many controls harbor undetected heart failure. This illustrates the narrow and diluted representation of clinical phenotypes by ICD codes, which could cause GWAS to overlook crucial SNPs and contribute to the missing heritability problem.

Phenotypic heterogeneity is usually caused by genetic and environmental differences. A concrete example of how this problem unfolds is the seemingly homogeneous diagnosis of atherosclerotic heart disease (AHD), which is a contributing factor to heart failure. An ICD10 code for AHD only suggests atherosclerosis, not the proportion of heritable calcified plaques or associated risk factors. Given that heart failure usually manifests with clear cardiac dysfunction [9], AHD might follow suit, presenting different cardiac comorbidities based on the underlying pathways. If AHD is primary, heart failure and atrial fibrillation are common [10, 11], while atrioventricular blocks can occur if diabetes is involved [12]. This complex interrelation underlines the need for a more informative phenotype representation in SNP discovery, hence the collective study of these diseases under the AHF umbrella.

GxE interactions also contribute to phenotypic heterogeneity when variation in genotypes’ effects on a phenotype depends on environmental factors. For example, smokers carrying the T allele for rs7178051 have a 12% reduced risk of CHD, while carriers who have smoked only have a 5% reduced risk [13]. The T allele is associated with reduced *ADAMTS7* mRNA expression levels, while smoking has been associated with upregulation. This suggests that smoking reverses the T allele’s protective effect without introducing additional risk to non-carriers, though it is not known why suppression of *ADAMTS7* expression is cardioprotective. This is an example of how GxE interactions worsen phenotypic heterogeneity because the T allele for rs7178051 would explain less variance in CHD status if smoker status was ignored.

It is possible that GxE interactions worsen phenotypic heterogeneity, while phenotypic heterogeneity simultaneously makes GxE interactions more difficult to detect. To illustrate the former case, consider that twins living in the same household share far more environmental factors than unrelated GWAS participants, which makes GxE interactions and their contributions to heterogeneity difficult to detect with GWAS. To illustrate the latter case, note that GxE interactions will be diluted in GWAS if they only influence a subset of homogeneous phenotypes that comprise an observed trait. For example, consider two genetically distinct heart conditions, A and B, which manifest as AHF. They are phenotypically indistinguishable, so patients with A and/or B would be labeled as cases, and patients with neither as controls. Now suppose that a pure GxE interaction affects A but not B. In a twin study, monozygotic twins are more likely to share all heritable conditions than dizygotic twins regardless of differences in their genetic architecture, so the measured heritability of AHF would be the weighted average of the heritabilities of A and B. However, B’s presence in the GWAS would dilute the ability of investigators to detect the GxE interaction effect on A because B is not influenced by it, and encoding A and B together as AHF assumes that GxE interactions influence A and B similarly. Therefore, the simultaneous consideration of phenotypic heterogeneity and GxE interactions may be essential to understand AHF’s complete genetic architecture. The TRACE method we present here addresses these issues.

The first step of TRACE is to create relatively homogeneous AHF-related phenotypes and analyze them with GWAS. Pleiotropy studies repeatedly illustrate the interdependence of correlations between genotypes and phenotypes [14], which also describes individual ICD codes. However, we hypothesize that the latent dimensions of sufficiently many AHF-related ICD10 codes correspond to homogeneous unobserved phenotypes that will correlate to genotypes more robustly. We call these constructs ‘latent phenotypes’, which are determined by genotypes and the environment, and in turn give rise to the observed ICD10 codes. This **many genotypes → few latent phenotypes → many ICD10 codes** relationship requires fewer connections than the traditional Pleiotropy model of **many genotypes → many ICD10 codes**, making it more parsimonious (Figure 1a). Figure 1b illustrates how latent phenotypes can be computed from multiple ICD10 codes per person. From a practical perspective, pooling information from ICD10 codes will offset the noise and bias of each one and add context to the AHF-related phenotype.

**Figure 1.**
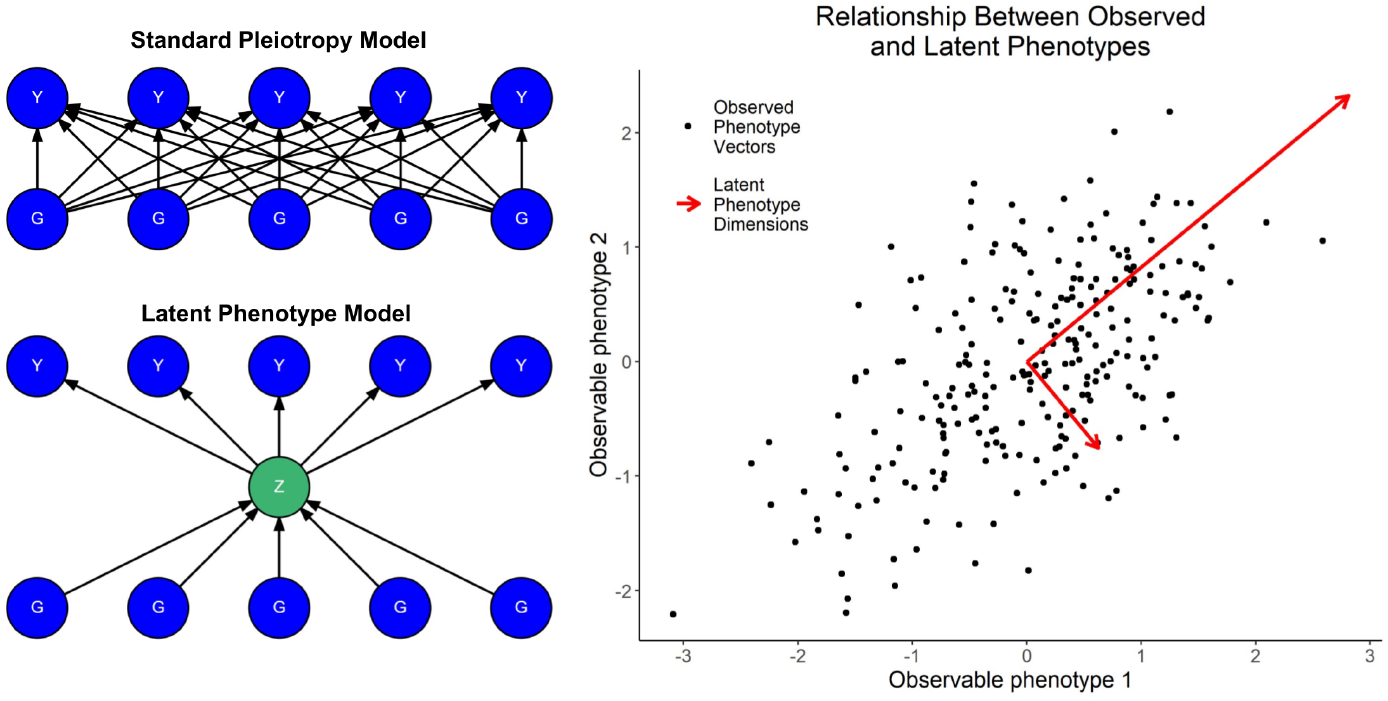
Schematic representation of our hypothesis and its operationalization. **[a]** Bayesian networks illustrating our hypothesis. In the presence of biological pleiotropy, latent phenotypes (Z) act as mediators, simplifying the genotype (G) to phenotype (Y) relationship. Only one latent phenotype and ten related variables out of many possible ones are shown for simplicity. **[b]** Each point on the scatter plot represents a phenotype vector. Suitable dimensionality reduction, such as PCA, could yield latent phenotype axes (shown as arrows). The figure represents observed phenotypes as continuous for illustrative purposes, showing how dimensionality reduction of many ICD codes might define a latent phenotype space, potentially offering a more informative phenotype representation and more robust genotype correlations”

After transforming ICD10 codes into latent phenotypes, TRACE’s next step is to uncover GxE interactions affecting them. If GxE interactions are diluted by phenotypic heterogeneity, and if latent phenotypes are more homogenous than ICD10 codes, then we expect to discover more GxE effects on our latent phenotypes. Additionally. most GWAS studies assume additive GxE effects, which means that the product of an additively encoded SNP and environmental factor is included in the GWAS regression. Our TRACE method challenges this oversimplification by incorporating categorical main and GxE effects into each SNP’s encoding prior to linear regression against the phenotype. This encoding is based loosely on the EDGE method [15], which essentially uses target encoding [16] to incorporate categorical genetic effects into each SNP’s encoding. However (letting T refer to an arbitrary test statistic), while target encoding generally substitutes *x* = *T*(*y*|*x*), our TRACE encoding generalizes *T*(*y*|*x*) to *T*(*y*|*x, E*), where *E* is an environmental factor. Thus, searching for GxE effects on latent phenotypes with TRACE may provide crucial insights into AHF’s heritability.

## Methods

### Transformative Regression Analysis of Combined Effects (TRACE)

Figure 2 showed how our TRACE method detected pure GxE effects. It first detected combined main-GxE effect by encoding both effects into SNPs and regressing them against latent phenotypes. Each SNP-environmental factor pair was encoded as a six-column matrix. Columns one and two were binary features with a value of 1 if the SNP had one or two minor allele copies, respectively, and a value of 0 otherwise. Columns three and four were obtained by multiplying columns one and two by the environmental factor column. Column five was the environmental factor, and column six was a constant. Multivariable linear regression was performed for each SNP against each latent phenotype using these columns, and the resulting SNP-informed beta coefficients {β_1_, β_2_, β_1*xE*_, β_2*xE*_} were used to compute the new SNP encodings. SNPs with {0, 1, 2} minor allele copies were encoded as {0, β_1_ + β_1*xE*_*E*_*i*_, β_2_ + β_2*xE*_*E*_*i*_} respectively, where *E*_*i*_ represented the *i*^*th*^ patient’s environmental factor value. SNPs with fewer than 1000 homozygous minor instances were binarized ({0, 1, 2} → {0, 1, 1}) prior to computing their TRACE encodings.

**Figure 2.**
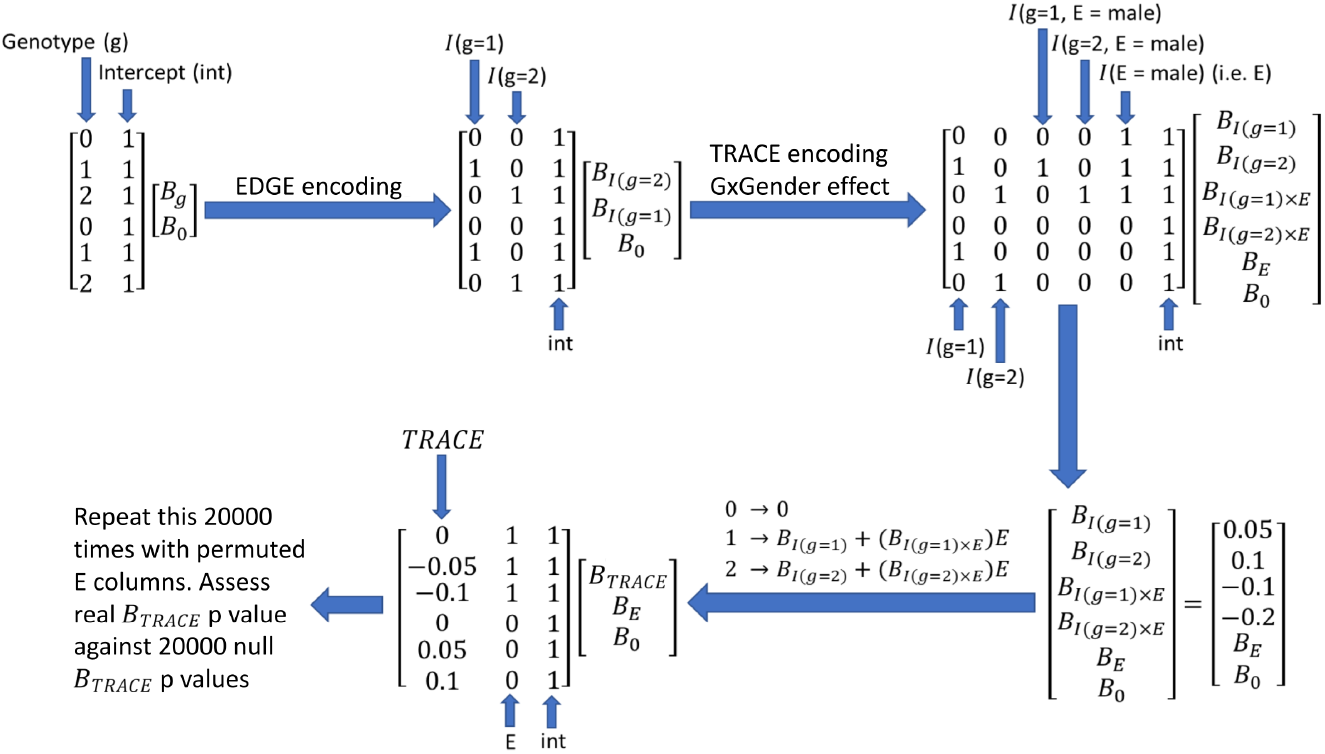
Figure 2: Schematic illustration of the TRACE method for detecting GxE effects. The diagram shows the SNP encoding process, beginning with a single additive genotype encoding (upper left), progressing through the initial EDGE transformation (upper middle), and culminating in the TRACE transformation (lower middle). In the final TRACE transformation, heterozygous additive SNP encodings are replaced with β_1_ + β_1*xE*_*E*_*i*_, and homozygous minor additive encodings with β_2_ + β_2*xE*_*E*_*i*_. Here, *E*_*i*_ represents the environmental factor (specifically gender in this example) for the *i*^*th*^ individual. Hypothetical beta coefficient values of {β_1_, β_2_, β_1*xE*_, β_2*xE*_} = {0.05, 0.1, –0.1, –0.2} are included for illustrative purposes, noting that TRACE computes them by regressing the 6 columns against a latent phenotype.

The main model (*H*_*alt*_) linearly regressed each latent phenotype against each TRACE-encoded SNP, its environmental factor covariate, and a constant. A null model (*H*_*null*_), containing only the environmental factor and constant, was also regressed against each phenotype. Nominal p-values for combined significance of main and GxE effects were determined with a likelihood ratio test statistic (λ) between the two models. More precisely, Nominal p-values were computed as follows.

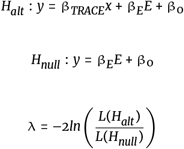

Here, *y* was a latent phenotype, *x* was a TRACE-encoded SNP, and *L* was a model’s likelihood function. The final step was to compute GxE effects by assessing each combined effect against a distribution of null effects with permuted environmental factor columns as described in the GWAS: GxE Effects section.

### Study Population and Genetic Data

Our study population included all UKB participants of self-declared white European ancestry, which was defined as having a coding 1, 1001, 2001, or 3001 for data field 21000 [17]. Individuals with heterotrophic cardiomyopathy were removed from the dataset as described by Aragam and colleagues [7]. We used Plink [18, 19] to remove SNPs from the dataset if they had a Hardy-Weinberg equilibrium p-value less than 10^−6^ or if they had a missingness rate exceeding 0.02. We then used Plink to remove individuals from the dataset if they had an average SNP missingness rate exceeding 0.02, if their SNP heterozygosity was more than 3 standard deviations from the mean, or if their X chromosome heterozygosity did not clearly indicate a binary sex chromosomal pattern. We used KING [20] to list all pairs of individuals with a kinship coefficient exceeding 0.0442. We then pruned individuals from the dataset to ensure that all pairs had kinship coefficients less than 0.0442 in a way that retained as many AHF cases as possible. We used Eigensoft [21, 22] to compute the top 20 PCs from a subset of SNPs that were thresholded with Plink to ensure a maximum SNP pair R2 of 0.09. The resulting dataset contained 14,762 AHF cases and 363,112 non-AHF cases, which were used as controls.

### Determination of Latent Phenotypes

ICD codes were selected from the set of all cardiovascular ICD10 codes (i.e., beginning with the letter I), select respiratory ICD codes (i.e., beginning with the letter J) that were known to be associated with heart failure, and the set of all miscellaneous ICD codes (beginning with the letter R). ICD codes that were not significantly correlated to AHF after a Holm-Bonferroni FPR correction were excluded from the latent phenotype generation. The remaining 311 ICD codes (table S1) and AHF were converted into three sets of 15 latent phenotypes; first by PCA, then logistic PCA, and finally an autoencoder. The autoencoder’s test-set reconstruction accuracy for the ICD codes was computed with 5-fold cross validation. For PCA and logistic PCA, a 16th latent phenotype was computed by estimating AHF from the first 15 latent phenotypes, and the estimated binary values’ residuals comprised the 16th latent phenotype. The autoencoder’s reconstruction error for AHF was less than 0.01%, which we believed was not enough variance to comprise a 16th latent phenotype, thus making the total number of phenotypes 47. All latent phenotypes were linearly regressed against the top 20 genetic PCs, and the Yeo-Johnson transformed residuals comprised the final latent phenotypes.

### GWAS: Main Effects

GWAS was conducted on the entire dataset. We computed main effect p-values using ordinary linear regression, with each SNP additively encoded and regressed against each latent phenotype. As demonstrated by the distribution of QQ-plot genomic inflation factors (Figure S1), they were less than or near 1.1 for all but two of the latent phenotypes. We deemed both main and GxE SNP hits statistically significant if their p-values fell below an adjusted genomewide threshold [23]. This adjustment divided the standard genomewide correction factor of 5 *×* 10^−8^ by 5*m, where m represented the ICC-corrected number of phenotypes, and 5 accounted for the number of effects tested. These effects included main effects and SNP interaction effects with smoking, alcohol, sex, and exercise.

### GWAS: Environmental Data

Our GxE interaction analysis focused on environmental factors: smoking, alcohol, sex, and exercise. Smoking was quantified using the UKB’s pack years of smoking feature (data-field 20161). Never-smokers (data-field 20160) were assigned zero pack years, and we imputed the remaining missing values. We calculated annual alcohol consumption by summing values from specific UKB data-fields (1568, 1578, 1588, 1598, 1608, 5364) and multiplying by the number of weeks per year. In the event of missing data, we resorted to the sum of corresponding monthly quantity data-fields (4407, 4418, 4429, 4440, 4451, 4462) multiplied by 12. If all contributing datafields were missing, or if any were declined (i.e. assigned values of -1 or -3), then we designated the annual alcohol consumption value as missing. Non-drinkers (data-field 1558) were assigned zero annual consumption, and we imputed the remaining missing values. Sex used a binary coding (1 for male, 0 for female). Exercise was represented as the first principal component of data-fields (874, 894, 914), post missing value imputation. Missing values within our modified alcohol and smoking features, and in datafields (874, 894, 914), were imputed with MICE [24, 25], which drew upon a range of other data-fields (table S2). MICE generally outperformed mean imputation, as indicated by simulations of random and nonrandom missingness (Table S3), unless frequent walkers were disproportionately absent in responses to data-field 874.

### GWAS: GxE Effects

Our TRACE method overestimated the significance of combined main-GxE effects because target encoding methods are known to overfit data [16]. To address this, a permutation test assessed the pure GxE effect for each SNP with a nominally significant TRACE p-value (the test statistic). For each nominally significant TRACE p-value, we computed 20,000 null p-values by permuting the environmental factor and recalculating the TRACE p-value, creating a null distribution that we would have expected in the absence of GxE effects. However, this distribution was noisy due to the limited number of permutations included. We addressed this by resampling 10,000 bootstrapped null distributions (each also of size 20000), fitting a Tukey-gh model to each bootstrapped distribution, and calculating the p-value of the original test statistic relative to each fitted Tukey-gh model. This generated a reliable distribution of 10,000 true p-value estimates, from which we used the 95th percentile as each SNP’s final GxE effect p-value, which ensured a false positive rate of 0.05.

### Predictive Models for AHF

SNPs with corrected genome-wide significant additive or GxE effects were used to predict their respective latent phenotypes. The SNPs’ additive contribution to these estimates, which we referred to as weighted SNP sums, were used as input for both logistic regression (LR) and gradient boosting classification (GBC). LR was used without regularization to explain AHF in terms of the weighted SNP sums. GBC was compared to LR to assess whether or not nonlinearity can improve the model’s fit. All models were validated with nested cross validation (30 outer folds and 10 inner folds) to confirm that any gains relative to LR were not a result of overfitting that can occur with standard cross validation. To ensure balanced case-control ratios and minimize bias, a subset of controls equal to the number of cases were randomly selected for each cross validation fold.

### Ontological Enrichment Analysis

We performed several *in silico* analyses to validate our SNP hits. Many of our main effects were validated by using LDtrait [26] to identify previous GWAS SNP hits that were within 500 kilobases (KB) of our SNP hits and had an *R*^2^ of at least 0.8. We used FUMA [27] to list all genes within 300KB of our SNP hits. We also used MSigDB [28, 29, 30] with the corresponding msigdbr R package [31] to identify gene sets that were significantly enriched for genes within 300 KB of each effect type. Among the gene-by-smoking interaction SNP hits, most of the enriched gene sets corresponded to miRNA molecules or transcription factors that were known to regulate those genes, so we used miEAA [32] to find KEGG pathways that were enriched for those miRNA molecules. We further examined the genes containing the aforementioned miRNA linked SNP hits by finding known literature associations with cardiovascular disease via DisGeNET [33, 34, 35, 36, 37]. Specifically, we entered a double-colon separated list of genes, downloaded the summary of disease-gene associations, retained only the rows containing a “Cardiovascular Diseases” flag in the “Disease Class” column, and ranked the strength of evidence that each gene was associated with cardiovascular disease by their gda score sums. We used Genevestigator [38] to rank the strength of evidence that each gene was differentially expressed in response to cigarette smoke.

### Shapley Value Explanations

We used ICD10 codes’ Shapley value contributions to the latent phenotypes to explain two correlations between latent phenotypes and miRNA linked gene-by-smoking SNP hits that resided inside of genes. We selected two genes for this follow-up analysis. One gene was primarily linked to cardiovascular disease with no significant ties to smoking, while the other had a notable smoking association but no clear link to cardiovascular disease. We extracted a minimal set of ICD10 codes, whose summed mean absolute Shapley values constituted 85% of the total. We then selected the smallest subset from these codes whose Shapley value sums (across individuals) correlated significantly with the TRACE-encoded SNP. For each subset size, we computed nominal GxE p-value proxies for all subset sums, treating each subset sum as a latent phenotype. The p-value proxies were defined as the nominal TRACE p-value divided by the main effect p-value, which correlated strongly (r = 0.87) with our permutation p-values (figure S2) for the applicable latent phenotype models. Identifying the permutation p-value for the subset with the lowest nominal GxE p-value allowed us to highlight the smallest subset size with a permutation p-value less than 5 *×* 10^−8^ that also yielded recurring ICD10 codes among the nominal top 20 subset combinations. These recurring codes, cross-referenced with related literature on the respective genes, provided plausible biological mechanisms for these gene-smoking interactions.

## Results

### GWAS: Main and GxE Effects

Applying TRACE to AHF uncovered numerous main and GxE interaction effects on 47 latent phenotypes, as detailed in table S4. Several dozen ICD10 codes contributed to the majority of our latent phenotypes, which are detailed in figure S3, though they broadly include various heart diseases, arrhythmias, vascular dysfunctions, and respiratory conditions. In this setting, we identified 300 SNPs significantly associated with latent phenotypes through main effects. We also identified 1207 gene-by-smoking interactions, 8 gene-by-sex interactions, and 11 gene-by-alcohol interactions. Table 1a counted two categories of independent SNP hits: “known” hits, which were in linkage disequilibrium (LD) with previously reported AHF-related GWAS hits, and “novel” hits, which were not. While there were an appreciable number of novel main effects, the majority of the gene-by-smoking SNP hits were novel, underscoring TRACE’s capacity to expand the GWAS search space. Because only one gene-by-exercise effect was detected at rs4382520, we did not further explore this interaction. We observed a range of non-additive GxE effects, spanning from partial dominance to over-dominance (Figure S4), which standard GWAS analyses would typically miss. Table 1b reinforces this finding by showing how a simple linear model produced significantly higher average p-values. In contrast, when we excluded non-additive effects (termed “conditional” in Table 1b), the linear model only marginally outperformed TRACE’s p-values.

**Table 1.**
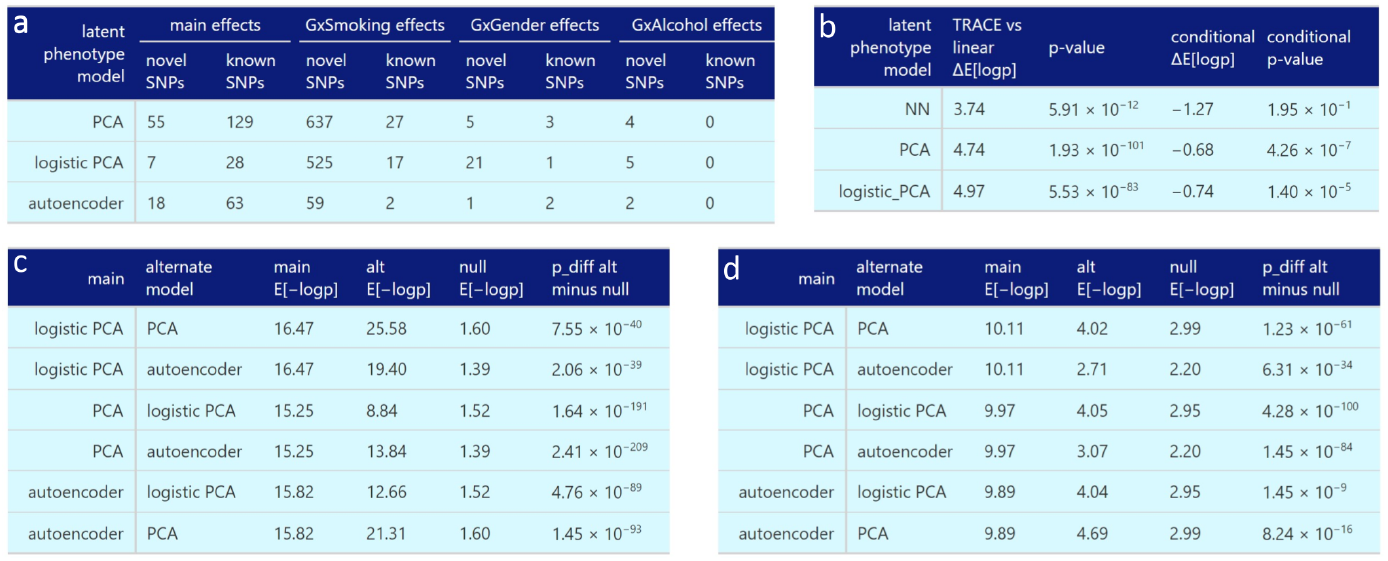
Tabulated counts of unique SNPs associated with all three latent phenotype models, followed by an aggregate comparison of their SNP hits. **[a]** Compares TRACE and linear regression in detecting nonlinear gene-by-smoking interactions, with TRACE improving by approximately 4 -log10p units. **[b]** Counts novel and known SNP associations across various effects, including main effects and interactions. **[c]** Cross-model analysis shows considerable overlap of significant main effect SNP hits, indicating that models capture largely overlapping biological information. **[d]** highlights that GxE interaction effects are more sensitive to latent phenotype construction, yet their average -log10 p-value proxy across models significantly exceeds random chance.

### Comparison of Main and GxE Effects

We assessed our latent phenotypes for their ability to capture similar biological information in the form of main and GxE interaction effects, which is important because they were derived from the same set of ICD10 codes. Table 1c quantified the SNP hit redundancy between models, demonstrating that each latent phenotype’s main effect SNP hits tended to maintain low average p-values in other models, significantly lower than what we would expect by chance. This confirmed that different latent phenotype models captured largely similar biological information, though not all main effect SNP hits achieved significance across all models, highlighting the importance of generating diverse latent phenotypes. Variance from GxE SNP hits was shared among latent phenotype models obtained for main effects via GWAS, but to a lesser extent. As shown in Table 1d, each model’s gene-by-smoking interaction SNP hits were associated with other models’ latent phenotypes at lower average p-value proxies (see the Shapley Value Explanations methods subsection) than expected by chance. However, these gene-by-smoking interaction SNP hits failed to maintain significance across different models more often than main effect SNPs, underscoring the heightened specificity with which GxE interaction effects contributed to latent phenotypes.

### Predictive Models for AHF

We used LR and GBC to predict AHF using weighted SNP sums derived from each model’s latent phenotypes. Figure 3a showed the *R*^2^ values between predicted AHF probabilities and actual AHF statuses with 95% confidence intervals (CIs). We compared LR and GBC on the same 30 validation folds for each model, and a paired Wilcoxon test confirmed that nonlinear GBC significantly outperformed LR in all three models. This finding aligned with recent evidence suggesting that human epistasis occurs at the systems level rather than the SNP level [39, 40]. Table S5 illustrated that the optimal complexity parameter (max tree depth) for GBC models remained consistent across outer validation folds, reducing the likelihood of overfitting or cross-validation bias in more complex models. Figure 3b compared our p-values to those mentioned in [7] for LR of SNPs against AHF. Noting that LR revealed only one significant SNP not reported by [7], figure 3b demonstrated the relative effectiveness of regressing SNPs against latent phenotypes.

**Figure 3.**
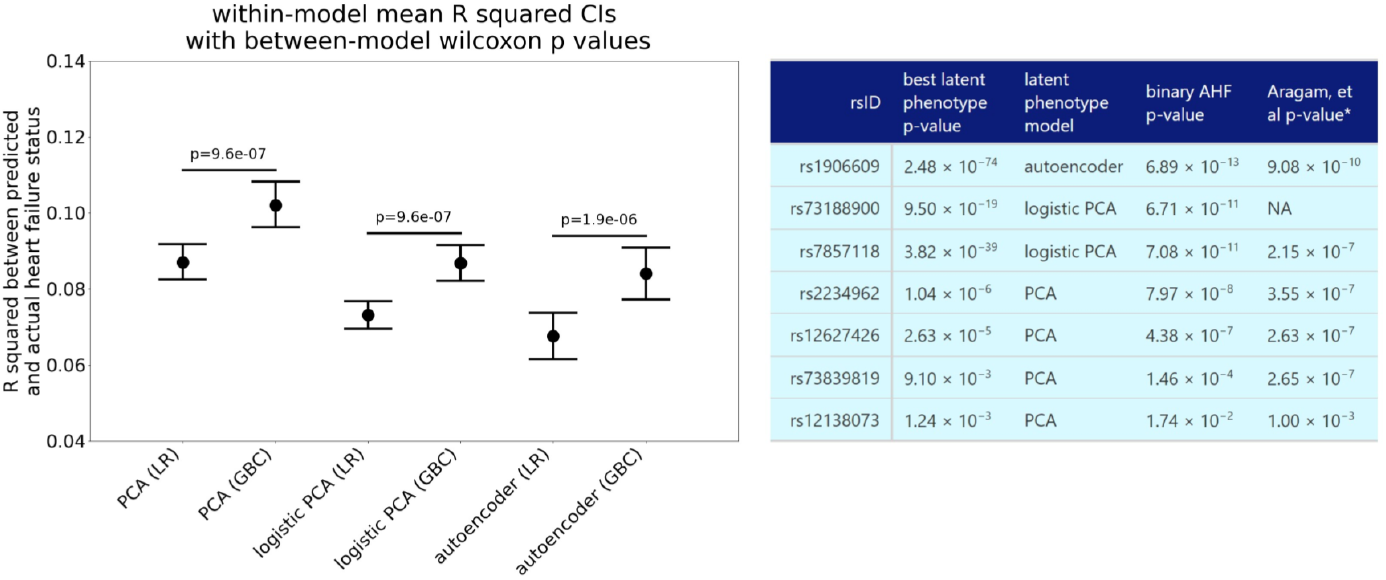
Performance comparison of predictive models for AHF using SNP-derived latent phenotypes. **[a]** Nested cross validation *R*^2^ values (with 95% bootstrapped CIs) illustrate the correlation between predicted and actual AHF statuses. GBC significantly outperforms LR across all models, as shown by paired Wilcoxon tests. **[b]** Comparative assessment of our latent phenotype p-values against LR (vs AHF) p-values and those from [7]. **[*]** The [7] p-value for rs12138073 could only be visually interpolated from their third figure, and is therefore a rough estimate.

### Ontological Enrichment Analysis

We examined the biological processes and pathways over-represented by genes in/near our SNP hits. Table 2a listed the top ten independent Gene Ontology (GO) pathways significantly enriched for genes within 300 KB of our main effect SNP hits. These pathways predominantly related to cardiovascular phenotypes, which substantiated the findings from table 1b. Table 2b listed the top ten KEGG pathways significantly enriched for miRNA molecules, where each miRNA molecule was enriched for genes within 300 KB of our gene-by-smoking interaction SNP hits. The top three pathways among these corresponded to growing evidence that terpenoids influence atherosclerosis [41], G protein-coupled odorant receptors regulate myocardial contractility [42], and smoking-induced impairment of ether lipid metabolism via LDL receptors in hepatocytes impacts cardiovascular phenotypes [43, 44, 45]. Table 2c listed the top GO pathways significantly enriched for transcription factor genes, each transcription factor being enriched for genes within 300 KB of our gene-by-sex interaction SNP hits. These pathways suggested a potential correspondence between our SNP hits and the enhanced vasculogenic potential of female hematopoietic stem cells [46].

**Table 2.** Examination of biological processes enriched for genes/miRNA linked to SNP hits against our latent phenotypes. **[a]** The top ten independent GO pathways significantly enriched for genes within 300 KB of our main effect SNP hits, emphasizing their key role in cardiovascular disease-related processes. **[b]** The top ten KEGG pathways significantly enriched for miRNA molecules, which in turn are enriched for genes in proximity to our gene-by-smoking interaction SNP hits. Several of these pathways suggest promising new avenues of research, including those related to terpenoid backbone biosynthesis (facilitates atherosclerosis in mice), olfactory transduction (regulates myocardial contractility), and ether lipid metabolism (regulates ion channels and correlates to cardiovascular health). **[c]** The most enriched pathways for transcription factor genes, which in turn are enriched for genes in proximity to our gene-by-gender interaction SNP hits. Note that our gene-by-gender interaction SNP hits enrich pathways related to both gender differentiation and cardiovascular disease, such as the heightened vasculogenic potential of female hematopoietic stem cells.

| a | GO gene set ID | Adjusted p-value | Number of genes |
| --- | --- | --- | --- |
| | gobp vascular process in circulatory system | $1.01 \times 10^{-5}$ | 13 |
| | hp cerebral inclusion bodies | $4.02 \times 10^{-4}$ | 6 |
| | gobp regulation of tube size | $1.70 \times 10^{-3}$ | 8 |
| | gobp regulation of blood circulation | $1.98 \times 10^{-3}$ | 10 |
| | gocc basal part of cell | $2.21 \times 10^{-3}$ | 10 |
| | gobp cell migration involved in heart development | $2.88 \times 10^{-3}$ | 4 |
| | hp neurofibrillary tangles | $2.88 \times 10^{-3}$ | 4 |
| | gobp regulation of blood pressure | $4.01 \times 10^{-3}$ | 8 |
| | gobp vascular transport | $4.01 \times 10^{-3}$ | 6 |
| | gobp heart process | $5.59 \times 10^{-3}$ | 9 |

| b | KEGG gene set ID | Adjusted p-value | Number of miRNA |
| --- | --- | --- | --- |
| | Terpenoid backbone biosynthesis | $4.78 \times 10^{-8}$ | 123 |
| | Olfactory transduction | $6.49 \times 10^{-7}$ | 256 |
| | Ether lipid metabolism | $1.37 \times 10^{-6}$ | 235 |
| | Nucleotide excision repair | $1.37 \times 10^{-6}$ | 159 |
| | Lipoic acid metabolism | $1.39 \times 10^{-6}$ | 15 |
| | Hippo signaling pathway - multiple species | $4.92 \times 10^{-6}$ | 188 |
| | Viral protein interaction with cytokine and cytokine receptor | $6.07 \times 10^{-6}$ | 234 |
| | Neuroactive ligand-receptor interaction | $9.95 \times 10^{-6}$ | 420 |
| | RNA polymerase | $1.97 \times 10^{-5}$ | 158 |
| | Various types of N-glycan biosynthesis | $2.05 \times 10^{-5}$ | 176 |

**Table 2.** Examination of biological processes enriched for genes/miRNA linked to SNP hits against our latent phenotypes.
| c | GO gene set ID | Adjusted p-value | Number of genes |
| --- | --- | --- | --- |
| | gland development | $4.13 \times 10^{-4}$ | 7 |
| | reproductive system development | $4.13 \times 10^{-4}$ | 7 |
| | sex differentiation | $4.13 \times 10^{-4}$ | 6 |
| | regionalization | $1.05 \times 10^{-3}$ | 6 |
| | columnar cuboidal epithelial cell differentiation | $1.05 \times 10^{-3}$ | 4 |
| | development of primary sexual characteristics | $1.63 \times 10^{-3}$ | 5 |
| | mononuclear cell differentiation | $2.00 \times 10^{-3}$ | 6 |
| | embryonic organ development | $2.00 \times 10^{-3}$ | 6 |
| | cell fate commitment | $2.00 \times 10^{-3}$ | 5 |
| | pattern specification process | $2.00 \times 10^{-3}$ | 6 |

Several pathways that extended beyond the top ten lists were also directly involved in cardiovascular disease. The top 11-25 gene-by-smoking interaction-linked miRNA KEGG pathways included renin secretion, ferroptosis, porphyrin and chlorophyll metabolism (integral to hemoglobin production), cholinergic synapses, arrhythmogenic right ventricular cardiomyopathy, cholesterol metabolism, and hypertrophic cardiomyopathy. A complete list of all enriched pathways from the gene set enrichment analysis were listed in Table S6. Despite the smaller number of gene-by-alcohol interaction SNP hits, genes within 300 KB of these hits were significantly enriched for few metabolic pathways. Each of these pathways, though enriched by only a single gene, was statistically significant. Furthermore, two out of the three associated genes had functions that could plausibly be impacted by ethanol.

Genes within 300 KB of our gene-by-smoking interaction SNP hits predominantly enriched gene sets regulated by specific miRNAs. We attempted to understand this observation with additional *in silico* analysis. For each latent phenotype model, a Fisher exact test confirmed that gene-by-smoking interaction SNPs were more likely to be within a gene if they were linked to an miRNA. This finding was congruent with the known regulatory roles of miRNAs at mRNA UTR regions and intronic sequences. We then cross-referenced these genes with external biological evidence. Table 3a listed the top five genes according to supporting study count, each exceeding a gda-score sum of 2 and an average differential expression (DE) ratio exceeding 1.5. While these genes broadly influenced various cell types, they consistently impacted cardiovascular disease-relevant pathways. In contrast, Table 3b presented the five genes with the highest gda-score sums and no appreciable DE in response to cigarette smoke. Note that four of these genes were almost exclusively expressed in tissues other than lung. Table 3c listed genes with the most supporting DE studies and a DE ratio exceeding 1.5, but without associations to cardiovascular disease, which suggested they are promising targets for future research. Finally, table 3d highlighted novel gene associations with no known links to cardiovascular disease or cigarette smoke exposure. For a complete list of miRNA linked gene-SNP pairs, see Table S7.

**Table 3.**
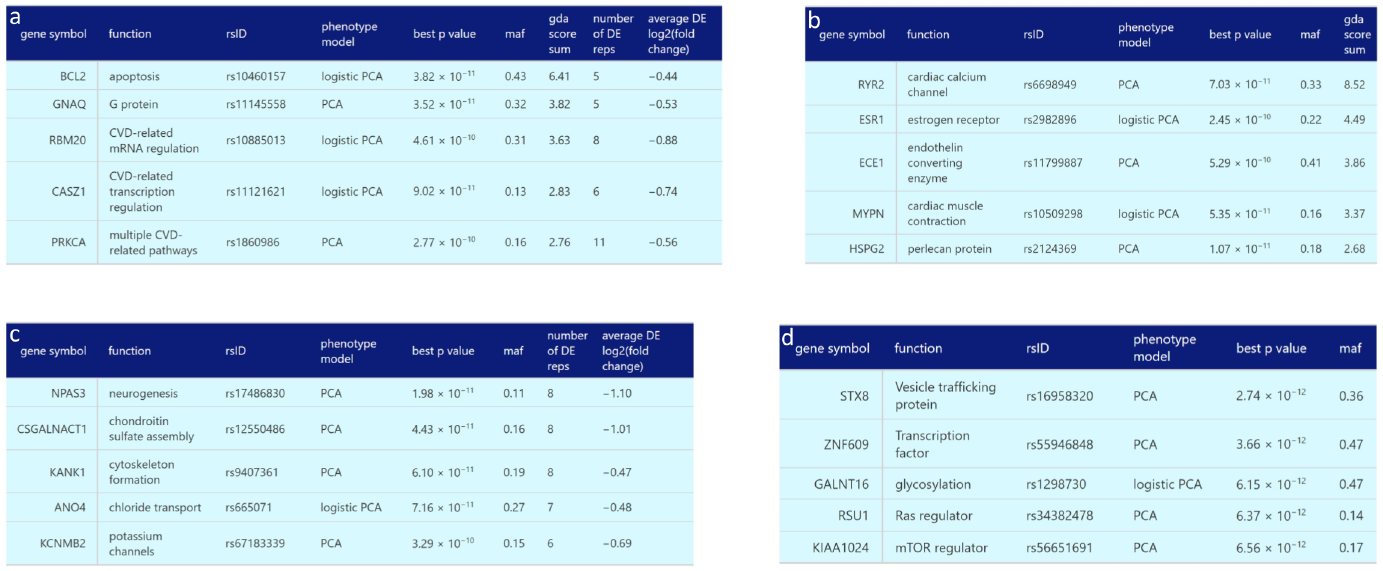
Detailed investigation of genes that contain a gene-by-smoking interaction SNP hit and are linked to a regulatory miRNA. **[a]** Top five genes, each with a gda-score sum over 2 and an average DE ratio exceeding 1.5, ranked by the number of supporting DE studies. **[b]** Five genes with the highest gda-score sums but lacking evidence of DE. Notably, *RYR2, ESR1, ECE1*, and *MYPN* are almost exclusively expressed in non-lung tissues. **[c]** Genes with the most supporting studies and a DE ratio over 1.5. These genes are not linked to cardiovascular disease, but the functions of *CSGALNACT1* [58], *KANK1* [59], and *KCNMB2* [60] are linked. **[d]** Top five genes (sorted by p-value) with no known links to cardiovascular disease or cigarette smoke. The functions of *STX8* [61], *GALNT16* [62], *RSU1* [63], and *KIAA1024* [64] are associated with cardiovascular disease.

### Shapley Value Explanations

Having analyzed the Shapley value contributions of ICD10 codes to latent phenotypes, we proposed potential biological mechanisms for two gene-by-smoking SNP effects. For the cardiac-specific *RYR2* gene in table 3b, subset analysis of Shapley values (size six) revealed that ICD codes I25.1 (atherosclerotic cardiomyopathy) and I25.9 (unspecified ischemic heart disease) were the most important contributors to the gene-by-smoking correlation between PCA latent phenotype 12 and rs6698949. Given the literature on calcium channel blockers mitigating cigarette-induced hypertension [47], a potential *RYR2*-smoking interaction could increase ischemic heart disease risk. We suspected this to occur via a non-atherosclerotic pathway because the Shapley values for I25.1 and I25.9 were anticorrelated. For the cigarette-related *KCNMB2* gene in table 3c, subset analysis of Shapley values (size 8) revealed that ICD10 code I83.9 (varicose veins) consistently contributed the most to the correlation between latent phenotype 12 and rs67183339. While numerous other ICD codes were prominent, we note that *KCNMB2* is a large conductance, voltage and calcium-sensitive potassium channel, and since these specific potassium channels mediate the link between Matrix metalloproteinase 2 and varicose veins [48], a *KCNMB2* SNP variant and cigarette smoke may interact to precipitate varicose veins. This aligned with findings that linked smoking to increased varicose veins risk [49, 50, 51], and varicose veins to heightened cardiovascular disease risk [52]. These proposed mechanisms provide novel hypotheses for further verification and study.

## Discussion

Most of the GxE effects detected were interactions with smoking, a finding that aligns with prior GxE effects that were identified for coronary artery disease and high blood pressure [53]. However, the effects identified in previous studies number around 40, whereas our approach found over 1000. The increased number of GxE effects and their consistency with prior literature’s effect types suggest that our original hypothesis is correct. Specifically, phenotypic heterogeneity dilutes GxE effects, and our latent phenotypes correspond to unobserved phenotypes that are more homogeneous. Unfortunately, our study is limited by the lack of replication of results, which is made difficult by the lack of a resource comparable to the UKB. However, future research could strengthen our findings by validating our GxE effects in an independent dataset. Our method could be applied to uncover novel genetic and GxE effects for any complex disorder, including depression, whose inherent heterogeneity has often resulted in SNP replication failure [54].

The identification of numerous non-additive gene-by-smoking interaction effects by TRACE is reminiscent of the considerable portion of heterosis in maize attributed to epistasis [55]. The possibility that epistasis contributes to AHF is further supported by recently discovered epistatic main effect contributions to cardiovascular disease [56]. While we did not discover any heterotic main effects, numerous gene-by-smoking interaction effects showed partial heterosis, hinting that cigarette smoke could influence AHF’s epistatic variance, consistent with the observed environmental dependency of bacterial epistatic effects [57]. Future research should explore gene-by-gene-by-environment interactions in heterotic GxE hits, which may elucidate the genetic-environmental interplay in AHF.

We addressed the tendency for target encoding to overfit by validating our GxE interactions with a permutation test, with uncertainty quantified by bootstrapping the distribution of permutation test null p-values. However, our choice of the 95th percentile as the final p-value might be overly conservative, potentially missing significant SNP hits. Table 1a confirmed that, for statistically significant additive GxE effects, linear regression’s average -log10(p) values were usually significantly higher than those of TRACE. However, it is crucial to note that even robust-looking parametric statistics may hide inherent variability, especially in the context of ultra-low p-values frequently found in GWAS. These are often sourced from data sparse tails of distributions, leading to hidden uncertainties. While TRACE’s p-values for GxE effects may appear stringent, their robustness is absent from more powerful parametric approaches. Future work developing a linear regression-based test for TRACE should bear in mind the need to account for such variance.

We identified hundreds of miRNA-linked gene-by-smoking interaction SNPs situated within genes, where many of these genes had known involvement with cardiovascular disease or smoking. The fact that our miRNA-linked SNPs disproportionately resided within a gene suggested a complex interaction network involving gene regulation by miRNAs. A cohort study in patients with diverse smoking histories may identify specific miRNA signatures associated with AHF and smoking. Functional validation of the findings via experimental studies such as reporter gene assays following *in vitro* knockdown/overexpression of miRNAs would also help to verify the influence of miRNAs and the genes they regulate on AHF.

While several promising research efforts tackle the environmental component of gene-by-smoking interactions, discovering and incorporating biologically causal SNPs into the analysis remains challenging. Traditional explanations point to the specific correlation between SNPs and causal SNPs in one LD pattern [65], which may not hold in another. However, this fails to account for the absence of independent GWAS in groups with varying LD patterns to find nearby SNPs with compensatory effects. We propose that phenotypic heterogeneity, particularly that arising from biases in ICD code assignment, might significantly contribute to these inconsistencies [6]. Differences in ICD code assignment are directly influenced by socioeconomic conditions [66], which also influenced exposure to environmental factors that contribute to heart disease and ischemia [67, 68]. Thus, in addition to reducing total phenotypic heterogeneity, our TRACE method could reduce differences in phenotypic heterogeneity between populations by shifting focus from heterogenous individual ICD10 codes to homogeneous ICD10 code patterns.

### Potential implications

Our findings contribute to the growing body of evidence that phenotypic heterogeneity dilutes GWAS SNP hits, and our novel GWAS method demonstrates one way to manage these issues. Application of TRACE to AHF resulted in hundreds of SNP hits, providing a different perspective from GWAS based on binary classification of cardiovascular disease ICD10 codes. The correlation we observed between SNPs and sums of specific Shapley value subsets highlights the complex nature of heart failure and how binary features failed to capture this complexity. The high number of heterotic gene-by-smoking interaction effects highlights the potential benefits of using a flexible model that does not impose the assumptions of standard SNP encodings. Interestingly, we observed enrichment for the miRNAs linked to genes near our SNP hits in crucial KEGG pathways, but not for the genes themselves, which indicates a significant role for miRNAs in heart failure’s genetic regulation. Thus, a complex interplay of genetic and gene-environment interaction effects seemingly drives AHF, which was only revealed by latent phenotypes.

## Availability of source code and requirements

- Project name: TRACE
- Project home page: our github repo
- Operating system: linux for the bash scripts
- Programming language: Python, R, bash
- Other requirements: see requirements.txt on our github repo
- License: MIT

## Data availability

The input data are available for other researchers via the UKB’s controlled access scheme [69]. The procedure to apply for access [70] requires registering with the UK Biobank and compiling an application form detailing:

- A summary of the planned research
- The UK Biobank data fields required for the project
- A description of derivatives (data, variables) generated by the project

In addition, several publicly available bio-informatics tools with associated databases were used in this study:

- Genevestigator: We used this to compare genes containing SNP hits to the genes’ DE in response to cigarette smoke. It can be accessed at https://genevestigator.com/.
- LDTrait tool: We used this to find established GWAS SNP hits in LD with those of our study. It can be accessed at https://ldlink.nih.gov/?tab=ldtrait.
- DisGeNET: This public platform’s gda-scores were used to quantify the evidence that genes containing some of our SNP hits are related to cardiovascular disease. It is available at https://www.disgenet.org/search.
- FUMA: We used this to select all genes within 300kb of our SNP hits. It is available at https://fuma.ctglab.nl/.
- MSigDB: We used this for our enrichment analyses. It can be accessed at https://www.gsea-msigdb.org/gsea/msigdb/index.jsp
- miEAA: We used this for our miRNA enrichment analysis. It is available at https://ccb-compute2.cs.uni-saarland.de/mieaa.

Access to these online resources is publicly available, but specific usage may require user registration. Please refer to each resource’s respective website for details on access, data use policies, and terms of service.

## List of abbreviations

AHF: All-Cause Heart Failure
AHD: atherosclerotic heart disease
CHD: coronary heart disease
CI: Confidence Interval
DE: Differential Expression
GBC: Gradient Boosting Classification
GO: Gene Ontology
GWAS: Genome-Wide Association Studies
GxE: gene-by-environment
ICC: Intraclass Correlation Coefficient
KB: kilobases
LD: Linkage Disequilibrium
LR: Logistic Regression
MICE: Multivariate Imputation by Chained Equations
PCA: Principal Component Analysis
SNP: single nucleotide polymorphism
TRACE: Transformative Regression Analysis of Combined Effects
UKB: United Kingdom Biobank

## Author’s Contributions

JTG and JHM conceived of and planned the study. JHM provided the oversight and funding. JTG performed the analyses and drafted the manuscript. All authors reviewed results, assisted with interpretation, and edited the manuscript

## Acknowledgements

The authors thank the participants of the UK Biobank study for making their data available for study.

