## supplemental_files for "Improving Genetic Association Studies with a Novel Methodology that Unveils the Hidden Complexity of All-Cause Heart Failure": medRxiv_supplemental_figures.pdf

*medRχiv*, 2023, 1–4

**doi: xx.xxxx/xxxx**
**academic preprint**
**RESEARCH**

RESEARCH

### Supplemental Figures:

### Improving Genetic Association Studies with a Novel Methodology that Unveils the Hidden Complexity of All-Cause Heart Failure

John T. Gregg[1], Blanca E. Himes[1], Folkert W. Asselbergs[2] and Jason H. Moore[3,*]

[1]Department of Biostatistics Epidemiology and Informatics, University of Pennsylvania, Philadelphia, PA, USA and [2]Institute of Cardiovascular Science, University College London, London, England and [3]Department of Computational Biomedicine, Cedars-Sinai Medical Center, Los Angeles, CA, USA

[*] **Correspondence address.** Jason H. Moore, PhD, FACMI, FIAHSI, FASA, Chair, Department of Computational Biomedicine, Cedars-Sinai, 700 N. San Vicente Blvd., Pacific Design Center Suite G540, West Hollywood CA 90069, USA. Tel: +1-310-423-3521;

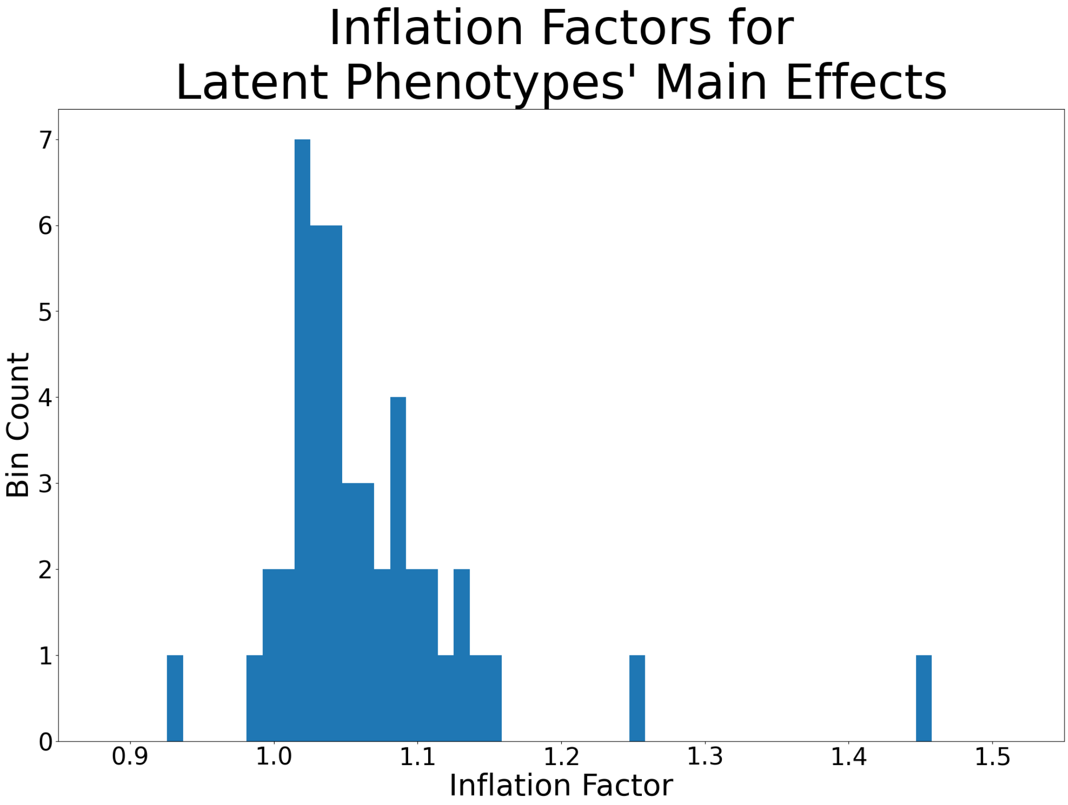


**Figure S1.** Distribution of genomic inflation factors for all latent phenotypes. The inflation factors are less than or close to 1.1 for all but two of the latent phenotypes, indicating a reasonable control of population stratification and/or cryptic relatedness in our analysis. Phenotypes with higher inflation factors were not further corrected because the majority of p-value inflation is caused by polygenicity [65], representing true biological signals. Note that LD score regression is not currently applicable to our TRACE method due to its multi-layered **genotype** $\longrightarrow$ **latent phenotype** $\longrightarrow$ **AHF** model.

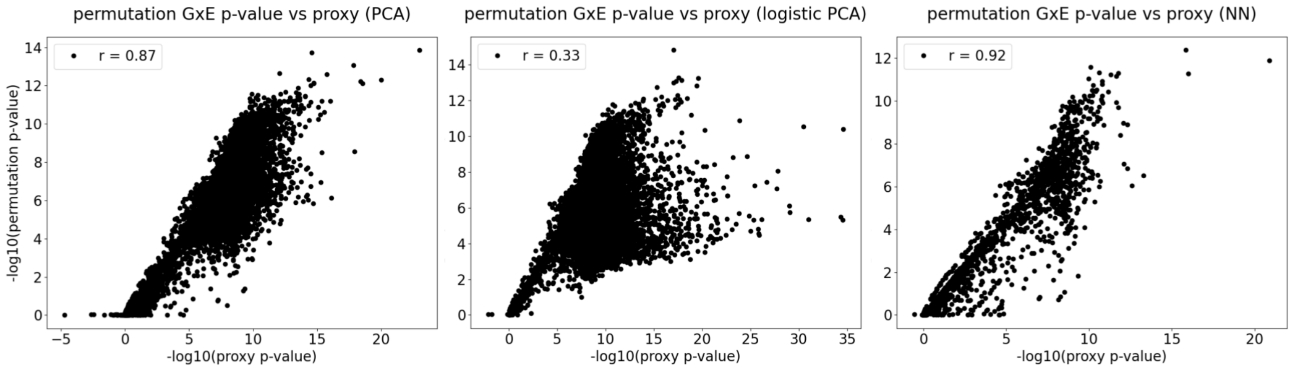


**Figure S2.** Correlation between nominal GxE p-value proxies and permutation p-values across all three latent phenotype models (S2a-c). The permutation GxE p-value proxies are computed as the nominal TRACE p-value divided by the main effect p-value for each latent phenotype. Across the three models, we observe strong correlations for PCA and the autoencoder (r = 0.87 and r = 0.92), but a weaker one for logistic PCA (r = 0.33). This discrepancy appears to be caused by increased inflation from the target encoding, which effects neither our use of the PCA p-value proxies nor the validity of the permutation p-values. Importantly, the p-value proxies were only used as an intermediate step to rank the correlations of Shapley value subset sums to specific genotypes. They were never used to assess statistical significance directly, so any potential biases and limitations do not impact our final results.

| ICD10 code | PCA average importance rank | Autoencoder average importance rank | Logistic PCA average importance rank |
|---|---|---|---|
| I10 | 1 | 3 | 2 |
| R07.4 | 3 | 6 | 1 |
| R11 | 4 | 7 | 5 |
| I25.1 | 2 | 12 | 7 |
| R31 | 7 | 10 | 4 |
| J90 | 17 | 4 | 8 |
| I25.9 | 8 | 8 | 15 |
| R69 | 6 | 9 | 20 |
| I20.9 | 5 | 14 | 19 |
| I84.9 | 15 | 11 | 13 |
| I48 | 9 | 27 | 3 |
| R55 | 10 | 23 | 6 |
| R33 | 11 | 21 | 9 |
| I25.2 | 18 | 2 | 22 |
| I48.9 | 13 | 17 | 12 |
| I95.9 | 23 | 5 | 16 |
| AHF | 12 | 13 | 21 |
| I83.9 | 16 | 16 | 17 |
| R07.3 | 14 | 32 | 10 |
| R13 | 20 | 29 | 11 |
| R42 | 24 | 24 | 14 |
| I51.7 | 27 | 1 | 47 |
| R51 | 21 | 40 | 18 |
| I26.9 | 29 | 35 | 23 |
| R32 | 42 | 18 | 29 |
| R06.0 | 19 | 47 | 24 |
| R91 | 41 | 26 | 27 |
| I25.8 | 26 | 36 | 38 |
| R53 | 31 | 39 | 31 |
| R00.0 | 43 | 15 | 43 |
| I21.9 | 30 | 33 | 40 |
| I50.1 | 22 | 56 | 25 |
| R00.2 | 34 | 48 | 26 |
| R05 | 33 | 49 | 30 |
| I50.0 | 25 | 43 | 45 |
| I95.1 | 46 | 41 | 36 |
| I84.8 | 63 | 28 | 34 |
| I47.1 | 54 | 19 | 52 |
| I21.4 | 40 | 45 | 41 |
| I73.9 | 44 | 37 | 46 |
| I21.1 | 51 | 22 | 56 |
| I20.0 | 35 | 44 | 50 |
| I48.0 | 37 | 58 | 35 |
| R35 | 50 | 52 | 28 |
| I63.9 | 52 | 30 | 53 |
| R00.1 | 38 | 60 | 39 |
| R07.2 | 32 | 64 | 42 |
| I45.1 | 56 | 25 | 58 |
| I51.8 | 36 | 20 | 87 |
| I84.2 | 60 | 50 | 33 |
| I35.0 | 48 | 38 | 62 |
| R15 | 57 | 59 | 32 |
| I80.2 | 47 | 65 | 37 |
| I50.9 | 28 | 88 | 48 |
| R21 | 61 | 72 | 44 |
| I21.0 | 55 | 68 | 60 |
| I34.0 | 39 | 55 | 92 |
| I67.8 | 58 | 61 | 67 |
| I69.4 | 73 | 54 | 61 |
| I47.2 | 71 | 34 | 86 |
| I42.0 | 75 | 53 | 68 |
| R18 | 53 | 89 | 55 |

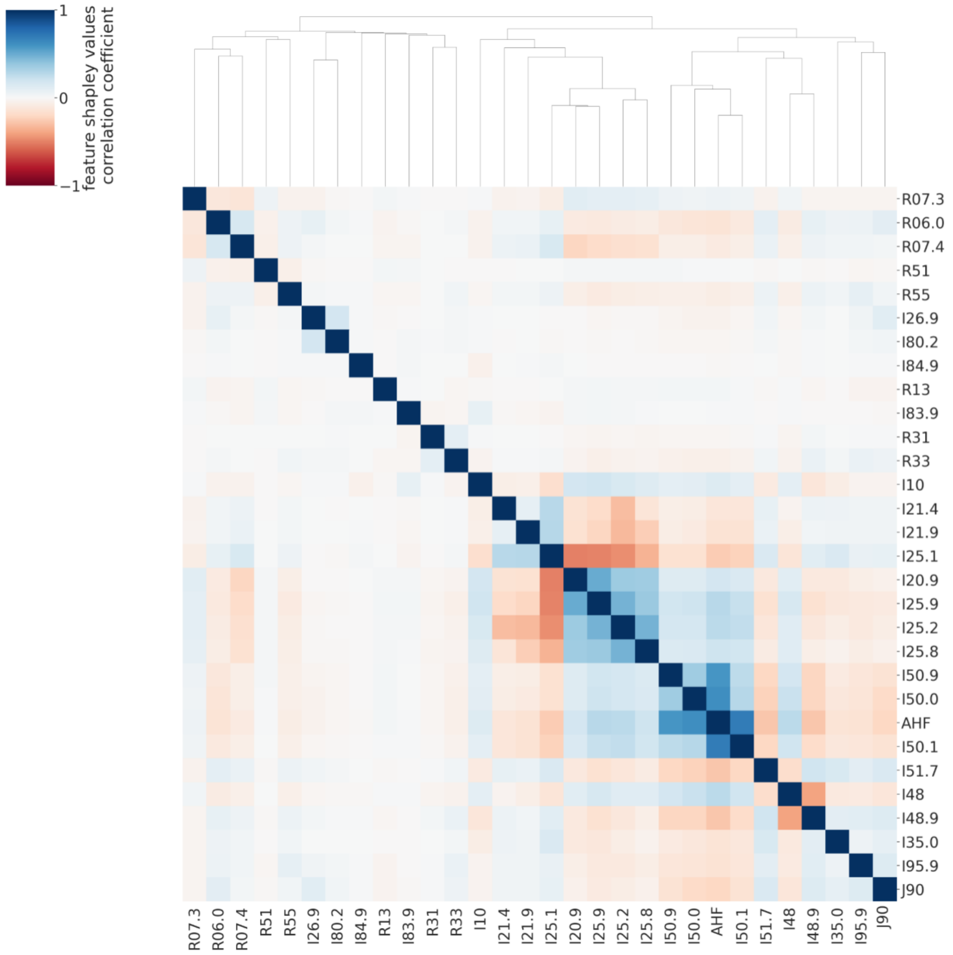


**Figure S3.** [S3a] ICD10 codes with the top 20% average feature importance ranks across latent phenotype models, which accounts for the majority of the total feature importance. For each ICD10 code, the respective importance rank in each model is provided. The Spearman $R^2$ values between the models' rankings are 0.38 for PCA vs autoencoder, 0,68 for PCA vs logistic PCA, and 0.31 for autoencoder vs logistic PCA. These correlations were only computed from the top 20% of features because the bottom 80% have minor importance, which artifactually increases the $R^2$ because the top 20% of features have consistently higher rankings. Loadings were used instead of Shapley values for Logistic PCA because inputting latent phenotypes are not computable by inputting ICD10 codes into a model, as is the case for PCA and autoencoders. [S3b] Heatmap represents Shapley value correlations for PCA latent phenotype index 12, which is selected for it's correspondance to the Shapley value analysis of two SNPs in table 3. The diagram illustrates the ways in which combinations of ICD codes cluster together, differentiating the latent phenotypes from one another. Key examples include the anticorrelated Shapley values for ICD10 codes I25.1 and I25.9, which significantly contribute to the correlation of the latent phenotype with rs6698949, and I83.9, which primarily contributes to the correlation with rs67183339. The fact that different subsets of Shapley values are primarily responsible for correlations to different genotypes underscores the complex genetic basis of All-Cause Heart Failure (AHF).

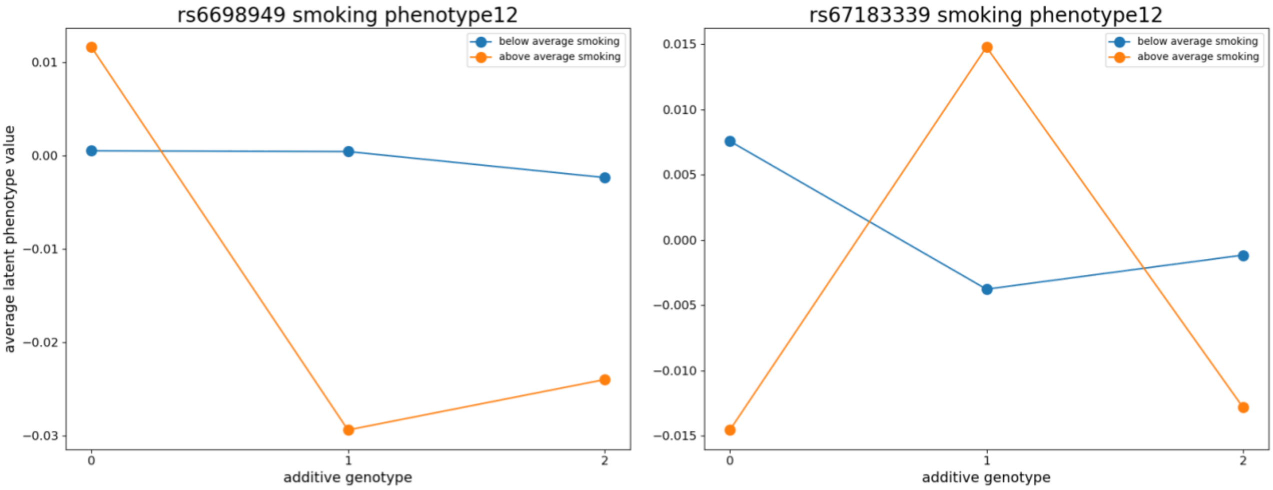


**Figure S4.** Illustration of non-additive Gene-Environment (GxE) interactions for two statistically significant SNPs linked to AHF. They were presented in table 3 and subsequently analyzed with Shapley value subset analysis, so their effects are supported by external biological evidence. The figures plot the average latent phenotype values for additively encoded genotypes {0, 1, 2}, further stratified by individuals who smoke at below and above average levels. The first rsID appears to demonstrate a dominance GxE effect, where heterozygous and homozygous minor individuals who smoke more than average have lower average phenotype values than individuals in the other four categories. The second rsID appears as a heterotic GxE effect with opposite directions for above and below average smokers. These non-additive GxE effects would be missed by standard GWAS.
